# Assessing AI tool use among New York State clinicians

**DOI:** 10.64898/2026.01.29.26345129

**Authors:** Andrew Galfano, Monica Barbosu, Beatrice Aladin, Ivelisse Rivera, Timothy D. Dye

## Abstract

Artificial intelligence (AI) is dramatically changing the healthcare landscape by providing patients, clinicians, administrators, and public health professionals with tools aiming to improve efficiency, outcomes, and experience in health. As elsewhere, New York State (NYS) experiences high demand for - and high investment in - transformation in healthcare with AI tools, though little is known about clinicians’ use and interest in adopting AI tools in their work. A large share of the nation’s future primary care clinicians train and work in NYS, and the state’s ability to establish clear policies, provide tools, and elevate AI competency have implications for care delivery nationally. As a result, we undertook this analysis of NYS clinicians’ use of AI to better understand opportunities for its adoption and inclusion in continuing education. For this analysis, we included healthcare providers who deliver ambulatory or specialty medical care within NYS, with use/frequency/purpose of AI tools by clinicians in their work as the main outcome. Of 305 NYS clinical providers responding, 23.4% indicated they use AI tools for work, and 11.1% report monthly use, 8.5% weekly use, and 4.6% daily use. AI was primarily used to search guidelines and ask clinical questions, followed by identifying drug interactions, analyzing data, analyzing images/labs, and creating care plans and patient recommendations. AI use did not vary significantly across professional disciplines or practice types, though independent practitioners were significantly more likely than advanced practice providers to use AI in their work, as were providers using social media and digital methods for obtaining continuing education. AI use increased substantially in 2025 compared with 2024. Overall, our findings suggest that programs targeting clinicians could incorporate these findings in designing accessible and acceptable AI-related continuing education opportunities to help familiarize clinicians with opportunities and risks for integrating AI tools into their practices.

**Author Summary:** AI tools are rapidly gaining traction in the delivery of healthcare. We found that clinician use of AI was quite limited (23%), though growing. Those using AI tools used them sparingly in their work, with only about 5% reporting daily use. The purposes for which clinicians report using AI – asking clinical questions, interpreting patient results, creating patient educational materials - could contribute substantially to healthcare outcomes if widely adopted. Designers of continuing education for clinicians should help provide opportunities for clinicians to improve their familiarity, use, and competency with AI tools, to help maximize the potential health benefits possible for patients and communities.

## Introduction

Artificial intelligence (AI) is dramatically changing the healthcare landscape by providing patients, clinicians, administrators, and public health professionals with tools aiming to improve efficiency, outcomes, and experience in health. Additionally, many consumers are using AI tools such as health-related chatbots to have their health questions answered. One in six adults regularly consults AI bots for medical information, reflecting that people may be less satisfied with their healthcare encounters, feeling that the wait times in offices are too long, doctors appear inattentive, and their healthcare bills are exponentially high.(Rosenbluth, 2025) People can quickly ask chatbots health-related questions without the need to contact a doctor. These experiences signal to the medical field that the status quo is under pressure to evolve, prompting the American Medical Association (AMA)’s recent call for AI in health care to be designed, developed and deployed in an ethical, equitable, transparent, and evidence-based manner.(American Medical Association, 2024)

Although AI technologies hold great promise for improving diagnosis, reducing administrative burdens, and offering personalized care, their uptake has been inconsistent, with major barriers spanning ethical concerns, workforce readiness, technological integration, regulation, and patient safety issues.(Ahmed et al., 2023) A substantial portion of healthcare practitioners still report limited usage, variable workflows, and lingering concerns about trust, oversight, and workflow fit.(Henry, 2025)

Studies across medical disciplines show that while practitioners generally express cautious optimism about AI, many report having limited formal exposure on how these systems work, contributing to hesitation.(American Medical Association, 2024) Heinrichs et al. found that clinicians tend to view AI as a potentially valuable aid, but vary widely in their comfort levels and knowledge.(Heinrichs et al., 2025) Broader research suggests that skepticism often arises from concerns about the accuracy of data, loss of clinical autonomy, fear of replacement, and the protection of patient privacy, especially among older practitioners who may have had fewer opportunities for digital training.(Heinrichs et al., 2025) At the same time, emerging studies indicate that integrated AI tools such as clinical documentation support in electronic medical records (EMRs) may help accelerate adoption by reducing the burden on clinicians and making AI use feel more natural within everyday practice.(Epic Systems Corporation, 2025) Together, these findings suggest that workforce training, ongoing education, and reliable access to tools are essential for realizing AI’s potential in healthcare.

New York State is uniquely positioned to shape the future of clinical AI, both due to its scale and the policy and innovation activities already underway across the state. For example, New York State has invested hundreds of millions of dollars in establishing “Empire A.I.,” reflecting the growing recognition that New York is moving toward a more coordinated approach and a national leadership role in AI adoption.(Ashford, 2024) At the same time, the state’s large and diverse clinical workforce makes this need especially urgent: New York State is ranked 8th in the nation for the most primary care providers per 100,000 population (United Health Foundation, 2025) and trains more medical residents and fellows than any other state, (Boyle, 2022) creating an opportunity to integrate AI into both clinical education and practice. With such a large share of the nation’s future primary care clinicians training and working in New York, the state’s ability to establish clear policies, provide tools, and elevate AI competency will have implications for care delivery nationally. As a result, we undertook this analysis of New York State clinicians’ use of AI to better understand opportunities for its adoption and inclusion in training programs.

## Methods

### Data source

We analyzed questions about AI use that were included in the Annual Needs Assessment of clinical providers in New York State (NYS), a survey developed for the NYS AIDS Institute’s Clinical Education Initiative (CEI; www.ceitraining.org). CEI aims to improve clinical care provided to people living with high-risk, medically complex conditions such as HIV/AIDS and HCV, as well as those needing sexual and drug user health services, by providing continuing medical education to healthcare providers. Advancing our understanding of how AI can be integrated into training is a priority. The Annual Assessment provides an opportunity to gauge clinicians’ learning priorities and modalities, and to ascertain trends in clinical care provision that could enhance the quality of care provided to the targeted affected communities. We used data from clinical provider assessments conducted in 2024 and 2025, which included questions regarding the use of AI in the workplace.

### Participants

For this analysis, we included healthcare providers who deliver ambulatory or specialty medical care within the State of New York. Providers are defined as those licensed medical professionals with prescribing privileges within New York State, as physicians (MD/DO), Physician Assistants (PA), Nurse Practitioners (NP), Certified Nurse Midwives (CNM), Dentists (DDS), and Doctors of Pharmacy/Registered Pharmacists (D/R PH).

### Sample selection

The assessment was deployed in English through the University of Rochester’s REDCap, a secure web application.(Harris et al., 2009) The survey link was widely distributed to the clinical provider community within New York State through the AIDS Institute’s Clinical Education Initiative newsletter, its website, social media, and email invitations to providers with accounts on the CEI learning portal.

### Measurement and data reduction

The primary outcome for this analysis is whether or not the provider used AI in their work:

> *How often do you use artificial intelligence tools for your work?*
>
> *Response categories: Never, Monthly, Weekly, Daily*

If the provider indicated that they used AI tools in their work on a monthly, weekly, or daily basis, we then asked: *For which of the following functions do you use AI tools?* and provided a checklist of potential options, including an open-ended response category.

The full distribution of the outcome variable is presented in Table 1. For analysis, due to the limited sample size of the individual categories, we created a binary variable (never use AI vs. use AI at least monthly) to simplify analysis and enhance the sample size of the “use AI” category.

**Table 1.** Frequency of AI Use at work with Functions of AI.

| **Table 1. Frequency of AI Use at work with Functions of AI** |  |
| --- | --- |
| **Variable** | **% (n)** |
| **How often do you use artificial intelligence tools for your work?** |  |
| Never | 75.7 (231) |
| Monthly | 11.1 (34) |
| Weekly | 8.5 (26) |
| Daily | 4.6 (14) |
| (No response) | 11 |
| Any use (at least monthly) | 23.4 (74) |
| Never use | 73.1 (231) |
| **Functions of AI tools:** |  |
| Ask clinical questions | 12.7 (40) |
| To search guidelines | 12.7 (40) |
| To identify drug interactions | 6.6 (21) |
| Analyze data | 6.0 (19) |
| Analyze images or labs | 5.1 (16) |
| To create care plans and recommendations for patients | 4.7 (15) |
| Other: | 5.1 (16) |
| “Charting - it's our new EMR platform” |  |
| “Create and revise documents, policies” |  |
| “Translation into other languages” |  |
| “EKG monitoring” |  |
| “Encounter notes” |  |
| “Expedite administrative work” |  |
| “Formulate arguments based on peer-reviewed data.” |  |
| “I use OpenEvidence like a librarian research assistant” |  |
| “Note writing with DAX” |  |
| “To create EFT scripts” |  |
| “To create lectures” |  |
| “To create policies” |  |
| “To reply to patient direct messages to provider” |  |
| “To write letters” |  |
| “To write recommendation letters, speeches, evaluations, etc.” |  |

### Statistical Methods

We used the Pearson chi-square test to establish associations between variables and the outcome measure of interest, defining statistical significance as the 2-sided asymptotic chi-square value, with significance <0.05. We included all variables in the multivariate model that were at least marginally associated with the outcome (p<0.25). We used the backward elimination logistic regression procedure to remove variables from the multivariate model (Wang & Cheng, 2020), with 0.25 as the cutoff p-value for entry into the model and 0.10 to remain in the model. (Bursac et al., 2008) We assessed the overall significance of the resulting multivariate model at p<0.05 and evaluated the model fit using the Pearson chi-square Goodness-of-Fit chi-square and p-value, an appropriately fit model was defined as p>0.05. We used adjusted Odds Ratios (aOR) to denote the same from multivariate analyses. We used SPSS 29.0.2.0 (IBM Corporation, Armock, NY, USA) for all analyses.

### Ethical review

The University of Rochester’s Research Subjects Review Board (RSRB) determined that this assessment meets federal and University criteria for exemption (STUDY00006285, RSRB00062100, and STUDY00006249). The project was conducted in accordance with the ethical standards outlined in the 1964 Declaration of Helsinki and its subsequent amendments.

## Results

Out of 305 clinical providers in New York State, 23.4% indicated that they have used AI tools for work (Table 1), with 11.1% reporting monthly use, 8.5% reporting weekly use, and 4.6% reporting daily use. AI was primarily used to search guidelines for medical care (12.7%) and ask clinical questions (12.7%), followed by identifying drug interactions (6.6%), analyzing data (6%), analyzing images or labs (5.1%), and creating care plans and recommendations for patients (4.7%).

As shown in Table 2, overall, AI use did not vary significantly across professional disciplines or practice types, although it was marginally more common in private practices compared to other practice types (p = 0.072). When grouped, independent practitioners (MD/DO/DDS/PharmD) were significantly more likely than advanced practice providers (PA/NP/CNM) to have used AI in their work (29.5% vs. 19.2%, p=0.036). No significant or marginal variation in AI use was observed based on provider specialization (data not shown). AI usage significantly increased, almost doubling from 2024 to 2025, rising from 18.3% in 2024 to 32.8% in 2025. Participants offering HIV screening to patients age 13 and over were less likely to use AI than those not screening for HIV, or than those who did not consider screening applicable to their context (Table 3). No other written institutional policies (such as PEP, PrEP, or STI screening) were related to the use of AI.

**Table 2.** Participant Characteristics Associated with AI Use.

| Characteristic | How often do you use artificial intelligence tools for your work? |  |  | p-value |
| --- | --- | --- | --- | --- |
|  | Total n | Never use AI n (%) | Use AI at least monthly n (%) |  |
| **What is your primary professional discipline/occupation?** |  |  |  | 0.107 |
| MD/DO | 102 | 75 (73.5) | 27 (26.5) |  |
| PA | 34 | 27 (79.4) | 7 (20.6) |  |
| NP/Nurse Midwife | 122 | 99 (81.1) | 23 (18.9) |  |
| DDS/PharmD | 47 | 30 (63.8) | 17 (36.2) |  |
| Independent practitioners (MD/DO/DDS/PharmD) | 149 | 105 (70.5) | 44 (29.5) | 0.036 |
| Advanced Practice Providers (PA/NP/CNM) | 156 | 126 (80.8) | 32 (19.2) |  |
| **In what setting do you perform medicine?** |  |  |  | 0.592 |
| Rural | 21 | 18 (85.7) | 3 (14.3) |  |
| Suburban | 20 | 15 (75) | 5 (25) |  |
| Urban | 262 | 196 (74.8) | 66 (25.2) |  |
| **AI use in 2024 vs 2025** |  |  |  | 0.004 |
| Use in 2024 | 180 | 147 (81.7) | 33 (18.3) |  |
| Use in 2025 | 125 | 84 (67.2) | 41 (32.8) |  |
| **Male vs Female** |  |  |  | 0.792 |
| Male | 87 | 65 (74.7) | 22 (25.3) |  |
| Female | 218 | 166 (76.1) | 52 (23.9) |  |
| **What is your primary practice setting?:** |  |  |  | 0.299 |
| CHC | 59 | 49 (83.1) | 10 (16.9) |  |
| Hospital or AMC | 55 | 41 (74.5) | 14 (25.5) |  |
| Private or Group Practice | 39 | 25 (64.1) | 14 (35.9) |  |
| Substance Use Center or Program | 28 | 22 (78.6) | 6 (21.4) |  |
| State Local Health Department Site | 24 | 19 (79.2) | 5 (20.8) |  |
| Correctional Facility | 13 | 10 (76.9) | 3 (23.1) |  |
| School or College | 26 | 22 (84.6) | 4 (15.4) |  |
| Retail/Commercial Pharmacy | 14 | 12 (85.7) | 2 (14.3) |  |
| Other | 46 | 230 (75.7) | 74 (24.3) |  |
| **Private Practice vs Other Practice Types** |  |  |  | 0.072 |

| **Characteristic** | **How often do you use artificial intelligence tools for your work?** |  |  | **p-value** |
| --- | --- | --- | --- | --- |
|  | **Total n** | **Never use AI n (%)** | **Use AI at least monthly n (%)** |  |
| Private Practice | 39 | 25 (64.1) | 14 (35.9) |  |
| Other Practice Types | 265 | 205 (77.4) | 60 (22.6) |  |

**Table 3.** Organizational Policies Associated with AI Use.

| How often do you use artificial intelligence tools for your work? |  |  |  |  |
| --- | --- | --- | --- | --- |
| Characteristic | n | Never use AI<br>n (%) | Use AI at least<br>monthly<br>n (%) | (p-value) |
| **Do you offer HIV tests to your patients 13 years and older?** |  |  |  |  |
|  |  |  |  | 0.035 |
| Yes | 213 | 170 (79.8) | 43 (20.2) |  |
| No | 33 | 22 (66.7) | 11 (33.3) |  |
| NA | 58 | 38 (66.5) | 20 (34.5) |  |
| **Participant's organization has written practice policies on the following:** |  |  |  |  |
| HIV Testing | 198 | 153 (77.3) | 45 (22.7) | 0.395 |
| HCV Testing | 158 | 122 (77.2) | 36 (22.8) | 0.533 |
| STI Screening | 173 | 131 (75.7) | 42 (24.3) | 0.994 |
| PrEP | 147 | 109 (74.1) | 38 (25.9) | 0.533 |
| PEP | 126 | 95 (75.4) | 31 (24.6) | 0.907 |
| Reassuring people that health information is not shared with ICE | 83 | 66(79.5) | 17 (20.5) | 0.346 |
| When and how to request language interpreter when serving patients whose primary language they do not speak | 183 | 141 (77) | 42 (23) | 0.513 |
| When and how to request an American Sign Language Interpreter | 130 | 98 (75.4) | 32 (24.6) | 0.901 |
| Health Equity | 120 | 90 (75) | 30 (25.0) | 0.809 |
| Screening for social determinants of health | 146 | 113 (77.4) | 33 (22.6) | 0.517 |
| Reproductive Health | 104 | 78 (75) | 26 (25) | 0.829 |

Overall, as shown in Table 4, the use of AI in practice was highest among individuals who preferred to participate in continuing medical education (CME) training through mobile apps (29.9%) and podcasts (30.2%), and lowest among those who preferred case-based training (22.3%). The use of AI was not significantly associated with other forms of CE training, such as online or in-person.

**Table 4.**
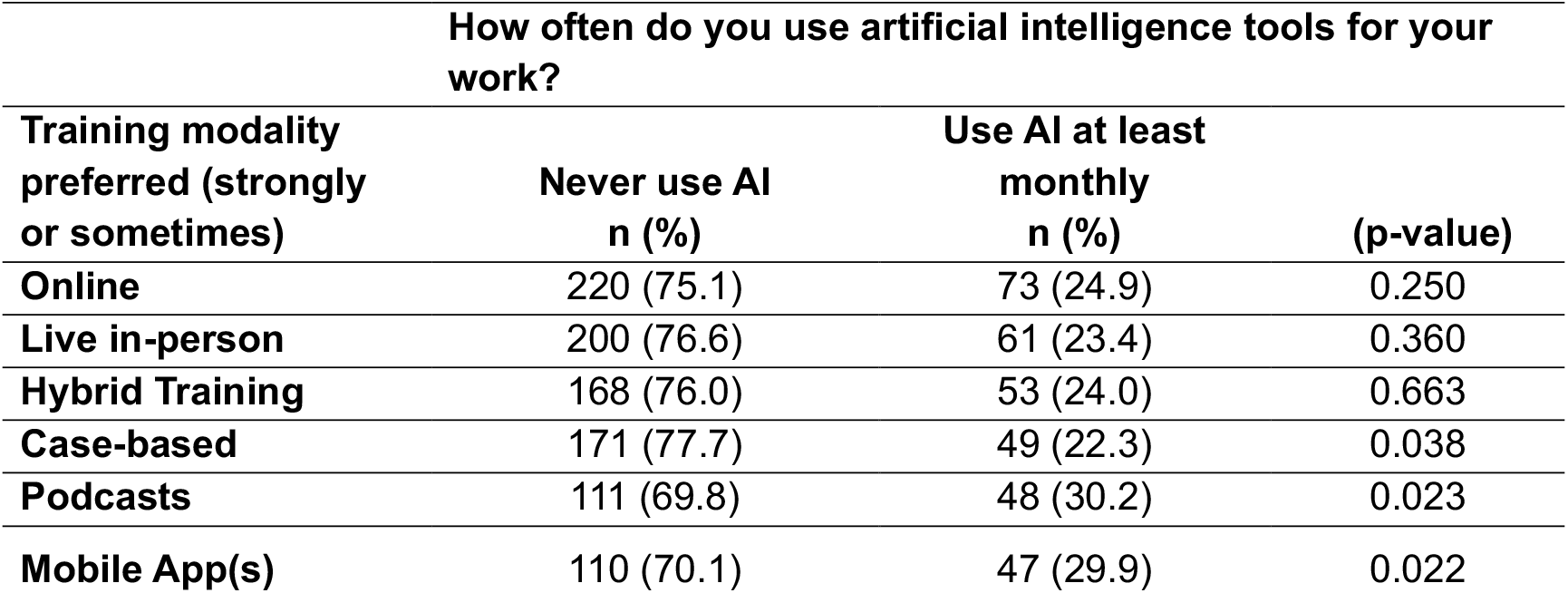
Preferred modalities for continuing education training and association with AI use in practice.

| **How often do you use artificial intelligence tools for your work?** |  |  |  |
| --- | --- | --- | --- |
| **Training modality preferred (strongly or sometimes)** | **Never use AI**<br>**n (%)** | **Use AI at least**<br>**monthly**<br>**n (%)** | **(p-value)** |
| **Online** | 220 (75.1) | 73 (24.9) | 0.250 |
| **Live in-person** | 200 (76.6) | 61 (23.4) | 0.360 |
| **Hybrid Training** | 168 (76.0) | 53 (24.0) | 0.663 |
| **Case-based** | 171 (77.7) | 49 (22.3) | 0.038 |
| **Podcasts** | 111 (69.8) | 48 (30.2) | 0.023 |
| **Mobile App(s)** | 110 (70.1) | 47 (29.9) | 0.022 |

Participants who used social media for work purposes were nearly twice as likely to use AI in their practice compared with those who did not use social media (29.7% vs. 14.5%; Table 5). Participants who used specific social media platforms, such as Facebook, Instagram, X (formerly Twitter), and YouTube, were significantly more likely to also use AI in their practices than were participants who did not use these platforms.

**Table 5.** Social media use for work purposes and AI use in practice.

| How often do you use artificial intelligence tools for your work? |  |  |  |
| --- | --- | --- | --- |
| Characteristic | Never use AI | Use AI at least | (p-value) |
|  | n (%) | monthly<br>n (%) |  |
| Social Media Use |  |  |  |
| No social media use | 94 (85.5) | 16 (14.5) | 0.003 |
| Yes social media use | 137 (70.3) | 58 (29.7) |  |
| Specific social media platforms |  |  |  |
| Facebook | 16 (57.1) | 12 (42.9) | 0.016 |
| LinkedIn | 54 (69.2) | 24 (30.8) | 0.120 |
| X (Formerly Twitter) | 4 (30.8) | 9 (69.2) | <0.001 |
| Instagram | 16 (51.6) | 15 (48.4) | <0.001 |
| WhatsApp | 19 (79.2) | 5 (20.8) | 0.683 |
| Youtube | 24 (57.1) | 18 (42.9) | 0.002 |
| Tiktok | 2 (50.0) | 2 (50.0) | 0.227 |

In multivariate analysis (Table 6), after controlling for other significant and marginal variables, independent practitioners and participants who preferred modern teaching modalities (e.g., podcasts, mobile apps) were more likely to use AI in their work, particularly in 2025 when AI use increased substantially compared with 2024. After controlling for confounders, the non-use of AI in the workplace was more common among clinicals who preferred more traditional learning modalities and among those who routinely offer HIV testing to their patients 13 years and older.

**Table 6.**
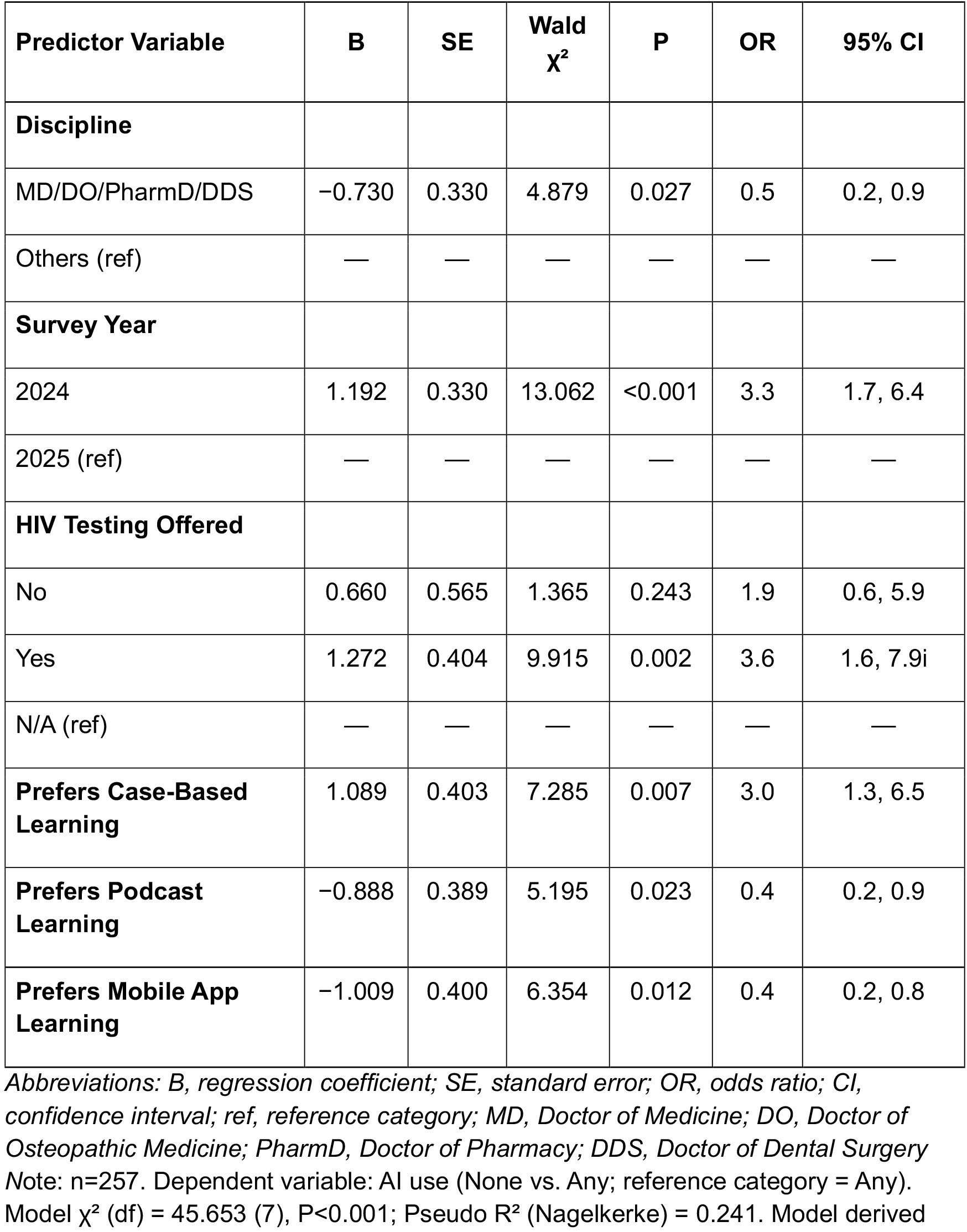
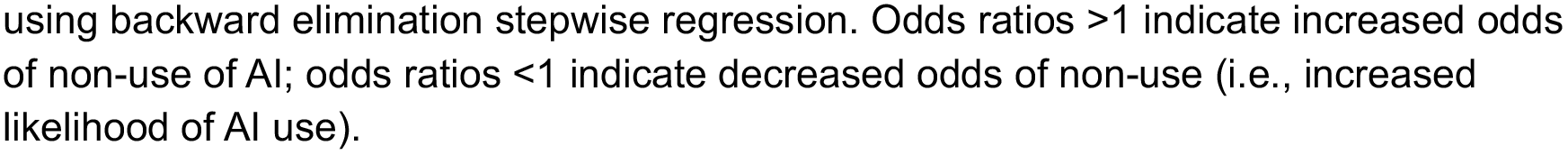
Multivariate analysis of Non-Use of Artificial Intelligence Among Health Providers, New York State.

| Predictor Variable | B | SE | Wald $\chi^2$ | P | OR | 95% CI |
| --- | --- | --- | --- | --- | --- | --- |
| **Discipline** |  |  |  |  |  |  |
| MD/DO/PharmD/DDS | -0.730 | 0.330 | 4.879 | 0.027 | 0.5 | 0.2, 0.9 |
| Others (ref) | — | — | — | — | — | — |
| **Survey Year** |  |  |  |  |  |  |
| 2024 | 1.192 | 0.330 | 13.062 | <0.001 | 3.3 | 1.7, 6.4 |
| 2025 (ref) | — | — | — | — | — | — |
| **HIV Testing Offered** |  |  |  |  |  |  |
| No | 0.660 | 0.565 | 1.365 | 0.243 | 1.9 | 0.6, 5.9 |
| Yes | 1.272 | 0.404 | 9.915 | 0.002 | 3.6 | 1.6, 7.9i |
| N/A (ref) | — | — | — | — | — | — |
| **Prefers Case-Based Learning** | 1.089 | 0.403 | 7.285 | 0.007 | 3.0 | 1.3, 6.5 |
| **Prefers Podcast Learning** | -0.888 | 0.389 | 5.195 | 0.023 | 0.4 | 0.2, 0.9 |
| **Prefers Mobile App Learning** | -1.009 | 0.400 | 6.354 | 0.012 | 0.4 | 0.2, 0.8 |

Abbreviations: B, regression coefficient; SE, standard error; OR, odds ratio; CI, confidence interval; ref, reference category; MD, Doctor of Medicine; DO, Doctor of Osteopathic Medicine; PharmD, Doctor of Pharmacy; DDS, Doctor of Dental Surgery Note: n=257. Dependent variable: AI use (None vs. Any; reference category = Any). Model $\chi^2$ (df) = 45.653 (7), P<0.001; Pseudo R<sup>2</sup> (Nagelkerke) = 0.241. Model derived
using backward elimination stepwise regression. Odds ratios $>1$ indicate increased odds of non-use of AI; odds ratios $<1$ indicate decreased odds of non-use (i.e., increased likelihood of AI use).

## Discussion

The American Medical Association (AMA) has stated that “AI has significant potential to advance clinical care, reduce administrative burdens, and improve clinician well-being.” The AMA has issued numerous policies regarding the development and use of AI in medicine, ensuring health equity, patient safety, and limiting risk to clinicians. (American Medical Association, 2024) Our study evaluated the usage of AI at work among various types of clinical practitioners in New York State. Our work provides insight into how practitioners are using AI, the purposes it serves, and how this use is concomitant with their use of other technology platforms such as social media, apps, and podcasts, and how these behaviors relate to workplace policies. Overall, we found that AI use remained relatively limited among New York State practitioners, with fewer than one in four reporting any use of AI in their work.

Though not widely studied across medical disciplines, AI is generally viewed positively in medical care but with cautious skepticism, (Albert, 2025) with male practitioners showing a slightly more positive attitude than female practitioners, and with chief physicians significantly less skeptical than residents. Heinrichs et al. explored attitudes toward AI, and found that most respondents viewed AI positively and considered AI to be an aid for medical staff.(Heinrichs et al., 2025) In a German study, Buck et al. explored attitude toward AI implemented in medical systems and found that clinical acceptance and adoption of AI balanced concerns and expectations, with unclear contribution toward AI attitudes of demographic and workplace characteristics. (Buck et al., 2022)

Other studies have consistently shown resistance to AI use in health care(Yang et al., 2024), which is similar to the results of our study, even with the substantial increase in reported AI use from 2024 to 2025. Overall, the use of AI among New York State clinicians in the workplace remained constrained, with no clear or consistently adopted workplace applications. Some researchers suggest that newer clinicians may be more receptive to AI use, helping to shape its role in the workplace, whereas more experienced clinicians, who are already comfortable in their practice, tend to be more cautious of new technologies and the associated challenges. (Shandilya & Fan, 2022) Although our study did not directly address the length of practice experience, higher AI use among clinicians who engage with technologies more commonly associated with newer practitioners, such as social media,(Shandilya & Fan, 2022) podcasts,(Schneider et al., 2024) and mobile apps.(Lee et al., 2023) suggest a similar trend. Further, some clinicians fear that AI could replace physicians or fail to understand the current practice guidelines, making the use of AI potentially more cumbersome than relying on clinicians’ own judgment.(Karches, 2018)

Our study revealed a significant increase in AI use in the workplace between 2024 and 2025, indicating potential shifts in the availability of AI tools. For instance, Epic’s AI tool, which transcribes clinical notes during patient visits, was released in 2025 (Epic, 2025). The increase in AI use in New York State could potentially be due, in part, to the growing availability of AI tools integrated into Electronic Medical Records (EMRs) systems.(Epic Systems Corporation, 2025) This could be due to corporations that are acquiring medical practices, requiring physicians to use these AI tools in their medical settings. Furthermore, EPIC and other EMR services are updating and integrating AI into their programs by default.

While this dataset offers valuable information on AI usage among various medical professionals, the study has several limitations. For example, our analysis is limited to providers working in New York State, and the use of AI may vary in other states due to different regulations and clinical guidelines. Currently, data on AI use within different medical specialty is limited, and we have not identified extensive examples of AI use, such as AI-assisted scribing, personalized targeted therapies, infection control in hospital, and support for complicated clinical decision making.(Haleem et al., 2019) Nevertheless, AI technology is already available, and everyday tools as search engines, computers and medical applications are adapting and implementing AI.

New York State clinicians are increasingly incorporating AI in their practice. Workplace policies and continuing medical education opportunities, especially those delivered through podcasts, social media, and apps, that support the effective and safe use of AI could enhance care for medically and socially complex patients, aligning with what AMA and other organizations envision.(American Medical Association, 2024) Clinicians have an opportunity to assess and enhance their AI-related knowledge and practical skills to improve patient care, expand access, and to manage the logistics and delivery of medical services. The Clinical Educating Initiative (CEI) can use the results of this study to create e-courses for how AI can be implemented into practices and facilitate teaching how to use AI tools to the participants.

## Data Availability

All data produced in the present study are available upon reasonable request to the authors

